# Subjective social status is associated with depressive symptoms beyond objective socioeconomic status among Chinese residents: a cross-sectional study

**DOI:** 10.64898/2026.04.30.26351164

**Authors:** Yiming Han, Tao Bo

## Abstract

**Purpose:** Socioeconomic status is strongly associated with depressive symptoms, but whether this reflects material conditions or perceived position is unclear. We tested whether subjective social status relates to symptoms independently of objective SES.

**Methods:** Using the 2022 Psychology and Behavior Investigation of Chinese Residents (PBICR2022), we analyzed 19,048 community residents. Objective SES was built by principal component analysis from income, education, employment, and housing. Subjective status was measured with a 7-point item and depressive symptoms with the Patient Health Questionnaire-9 (PHQ-9). Analyses used hierarchical regression, bootstrap mediation, moderated mediation, and response surface analysis.

**Results:** Probable depression prevalence was 21.77%. Subjective status was inversely associated with symptoms (= −0.098). Objective SES showed no overall association (total effect = −0.009, p = .30), with a weak protective association only among adults (18 years; −0.025, p = .004). Its protective value operated through subjective status: higher objective status went with higher identification, and higher identification with fewer symptoms (indirect effect = −0.020, 95% CI [−0.024, −0.016]). This channel was stronger in never-married than married adults (difference = −0.015, 95% CI [−0.026, −0.003]), though non-significant in stratified tests, thus suggestive. Never-married adults may lack this social validation and rely more on self-rated standing. No sex or migration differences appeared. Subjective status outweighed objective conditions, protection leveling off at high status.

**Conclusion:** Subjective status was more closely linked to symptoms than objective conditions, whose value operated mainly through raised subjective status. Cross-sectional design precludes causal inference.

## Introduction

A large body of research has found that higher social status is associated with better mental health [1–3]. However, whether what truly matters behind this association is objective material conditions or an individual’s subjective appraisal of their own social position remains unsettled. Objective socioeconomic status (SES) is usually indexed by income, education, employment, and housing, and it may protect against depression either by improving material conditions or by expanding life opportunities [1]. At the same time, subjective social status (SSS) refers to the self-positioning that individuals form through comparison with others. It translates objective circumstances into psychological experience and incorporates social comparison and judgments about fairness [4,5]. In Chinese samples, most studies have found that lower SSS is associated with more depressive symptoms, that this association persists after controlling for income and educational attainment [6,7], and that it is closer than that of objective SES indicators [3,8]. However, the overall association between objective SES and depression in Chinese populations is inconsistent, as previous studies have reported both protective and null associations [9–11].

Much of this literature has three limitations. First, most studies used regional or clinical samples, and few estimated these associations in a large national community sample with extensive covariate adjustment. Second, previous studies rarely stated explicitly whether objective SES remains meaningfully associated with depressive symptoms after controlling for subjective social status. Third, previous studies rarely tested whether objective status first affects subjective social status and then depression, and whether the strength of this chain is consistent across social groups. Existing comparisons typically split the sample into subgroups and estimated each separately without formally testing between-group differences, thereby ignoring sampling error [12].

In this study, we used data from the 2022 Psychology and Behavior Investigation of Chinese Residents (PBICR2022), a large national community sample, to examine four questions. (1) Is SSS still associated with depressive symptoms after controlling for objective SES and covariates? (2) Does objective SES show any meaningful overall association with depressive symptoms? (3) Does the indirect effect of objective status on depressive symptoms through subjective status differ by marital status, sex, or cross-provincial migration status? (4) Is the joint association of objective and subjective status with depressive symptoms non-linear, and is it dominated by subjective status? The third and fourth questions were tested with moderated mediation analysis and response surface analysis (RSA), respectively [13,14].

## Methods

### Study design and participants

Data came from the 2022 Psychology and Behavior Investigation of Chinese Residents (PBICR2022), a cross-sectional nationwide survey of community residents aged 12 years and older conducted through face-to-face and online questionnaires [15]. The survey used non-probability quota sampling, and respondents volunteered to take part. Because no sampling weights were provided, the results should be regarded as associational evidence from a large national community sample and cannot be generalized directly to the national population. Among 21,916 respondents, 2,867 (13.1%) who completed the questionnaire in less than 600 seconds were excluded, and one additional participant who reported an implausible subjective SES value was excluded, leaving a final sample of N = 19,048 (mean age = 39.01 years, SD = 18.74); details of the exclusion process are provided in the Supplementary Material.

### Measures

Depressive symptoms were assessed with the Patient Health Questionnaire-9 (PHQ-9; item 119 of the questionnaire) [16]. The scale contains 9 items with a two-week recall period, each rated from 0 (not at all) to 3 (nearly every day), yielding a total score from 0 to 27, where higher scores indicate more severe symptoms. A total score of 10 or higher was defined as probable depression [17].

Objective SES was constructed from four indicators: family per-capita monthly income (item 84, 10 levels), educational attainment (item 14, 9 levels), employment status (item 11, coded as employed), and housing conditions (items 81 and 82, housing area and number of dwellings, averaged). These four indicators represent economic resources, human capital, employment stability, and material wealth, respectively, and are the most commonly used dimensions for measuring objective social position [1]. Because the indicators differ in scale and are correlated, we extracted the first principal component by principal component analysis as the composite objective SES index [18]. This component explained 38.7% of the total variance. The index was standardized (M = 0, SD = 1), with higher values indicating higher objective status. The original response options and coding for each indicator are provided in the Supplementary Material.

Subjective social status (SSS) was measured with item 76 of the questionnaire, which asked how respondents would rate the social status of their family. Responses were scored from 1 to 7, with higher scores indicating a higher evaluation of one’s own family social standing [2].

### Covariates

The fully adjusted models controlled for age, sex, marital status (never married, married, divorced, widowed), family type, urban or rural residence, employment status (employed, not employed), number of chronic diseases, and province fixed effects. It should be noted that employment status both contributes to the objective SES index and is included as a covariate. With this treatment, the estimate for objective SES mainly reflects the roles of income, education, and housing. To check whether the results depend on this treatment, we report results from sensitivity analyses without the employment covariate.

### Statistical analysis

All continuous variables were standardized to a mean of 0 and a standard deviation of 1. The analyses first examined the reliability and structural validity of each measure, then described sample characteristics and correlations among variables, and subsequently examined the independent associations of subjective and objective status with depressive symptoms, the mediation of objective status through subjective status, group differences in this mediation, and the non-linear relationship between the two.

Hierarchical linear regression first added objective SES to the covariate model and then added subjective social status to examine their associations with depressive symptoms and the incremental variance explained by subjective status. Mediation analysis decomposed the association between objective status and depressive symptoms into an indirect effect through subjective social status and the remaining direct effect, with confidence intervals estimated using 3,000 percentile bootstrap resamples. Moderated mediation analysis compared the size of the indirect effect between men and women, between never-married and married adults (restricted to those aged 18 and older), and between local residents and cross-provincial migrants. Response surface analysis regressed depressive symptoms on objective status, subjective status, their squared terms, and their product term, with all covariates included.

To test robustness, we conducted six sensitivity analyses involving a reversed mediation model, different cutoff values, alternative SES construction methods, hierarchical R^2^ decomposition, and Harman’s single-factor test [19]; settings and results are provided in the Supplementary Material. Analyses were conducted in Python (pandas, NumPy, SciPy, scikit-learn, statsmodels).

In addition to the main analyses, we constructed a mismatch indicator defined as standardized subjective status minus standardized objective status, to directly examine whether the state of incongruence between objective conditions and subjective evaluation is itself associated with depressive symptoms. Two analyses were performed: first, the sample was divided into underestimation, matched, and overestimation groups by tertiles of the mismatch value to compare PHQ-9 differences; second, the mismatch value and objective status were entered into the model simultaneously to examine whether mismatch remains independently associated with depressive symptoms when objective conditions are held constant. Because the mismatch value is derived from the difference between subjective and objective status, entering all three into the same model would result in perfect collinearity; therefore, subjective status was not included in this model. In addition, a product term was included to test for interaction. These analyses were exploratory and were not corrected for multiple comparisons.

### Ethical considerations

The protocol was approved by the ethics committee of the Health Culture Research Center of Shaanxi (No. JKWH-2022-02), and all participants provided informed consent. Sex-based analyses and reporting followed the SAGER guidelines [20].

### Use of AI-assisted technologies

During the preparation of this work, the authors used DeepSeek, Kimi and ChatGPT to assist with the development of the data analysis code and to improve the readability of the manuscript language. After using these tools, the authors reviewed and edited the content as needed and take full responsibility for the content of the published article.

## Results

### Sample characteristics

Among the 19,048 community residents surveyed, the prevalence of probable depression (PHQ-9 10) was 21.77%. Table 1 summarizes the sample by sex. Women had a slightly higher mean PHQ-9 score than men (6.49 versus 6.33, p = .04), though the effect size was negligible; probable depression prevalence was identical between sexes (21.77% versus 21.77%, p = 1.00). Women’s objective SES was lower than men’s (−0.05 versus 0.06, p < .001), while subjective SES was similar (4.31 versus 4.33, p = .16).

**Table 1.** Sample characteristics by sex. Continuous variables compared by t-test; categorical variables by chi-square test.

| Variable | Overall (N = 19,048) | Female (n = 9,735) | Male (n = 9,313) | Statistic | p |
| --- | --- | --- | --- | --- | --- |
| Age (years), mean (SD) | 39.01 (18.74) | 38.73 (18.65) | 39.31 (18.84) | -2.101 | 0.04 |

| Variable | Overall (N =<br>19,048) | Female (n =<br>9,735) | Male (n =<br>9,313) | Statistic | p |
| --- | --- | --- | --- | --- | --- |
| Marital status, % |  |  |  |  |  |
| Never married | 39.6 | 38.7 | 40.6 | 7.534 | 0.006 |
| Married | 55.9 | 56.0 | 55.8 | 0.078 | 0.78 |
| Divorced | 1.8 | 1.9 | 1.8 | 0.373 | 0.54 |
| Widowed | 2.6 | 3.4 | 1.8 | 47.735 | < 0.001 |
| Urban residence, % | 69.9 | 69.6 | 70.3 | 0.995 | 0.32 |
| Cross-provincial migrant, % | 26.7 | 25.6 | 27.9 | 12.832 | < 0.001 |
| Objective SES (SD units), mean (SD) | 0.00 (1.00) | −0.05 (1.00) | 0.06 (1.00) | −7.591 | < 0.001 |
| Subjective SES (1-7), mean (SD) | 4.32 (1.29) | 4.31 (1.23) | 4.33 (1.35) | −1.398 | 0.16 |
| PHQ-9 score (0-27), mean (SD) | 6.41 (5.37) | 6.49 (5.16) | 6.33 (5.59) | 2.052 | 0.04 |
| Probable depression ( 10), % | 21.77 | 21.77 | 21.77 | 0.000 | 1.00 |

### Reliability and validity

The PHQ-9 showed good reliability and validity in this sample: internal consistency Cronbach’s = 0.912, KMO = 0.939, Bartlett’s test ^2^ = 91,054.28 (p < .001), and a single-factor solution explained 59.3% of the total variance, supporting its use as a unidimensional indicator of depressive symptoms. Subjective social status was measured with a single item, so internal consistency could not be computed, but its correlations with objective SES (r = 0.17) and depressive symptoms (r = −0.11) were both in the theoretically expected directions. Objective SES is a composite indicator; the four loadings were in the same direction, and the first principal component explained 38.7% of the total variance. Psychometric indicators are provided in the Supplementary Material.

### Associations of subjective social status and objective SES with depressive symptoms

The results of hierarchical regression are described below. The model containing covariates only explained 7.75% of the variance in PHQ-9 scores; adding subjective social status raised R^2^ to 8.58%, whereas adding objective status alone produced almost no change (7.76%). Of all incremental variance explained beyond the covariates, about 99% came from subjective social status, and the incremental contribution of objective status was minimal (ΔR^2^ = 0.0001). The fully adjusted response surface model explained 9.24%. In adjusted logistic models, objective SES was unrelated to probable depression at every cutoff. The conclusion remained unchanged after dropping the employment covariate. Therefore, in the total sample, no meaningful overall association existed between objective SES and depressive symptoms.

### Mediation analysis

As shown in Table 2, objective status was positively associated with subjective status, subjective status was negatively associated with depressive symptoms, and neither the total effect nor the direct effect of objective status was significant. The indirect effect transmitted through subjective status was small but statistically significant (Figure 1). This indicates that better objective conditions can raise the status individuals believe they hold, which in turn is associated with fewer depressive symptoms. The indirect effect was negative while the direct effect was positive in direction; the two run in opposite directions, so the total effect stays close to zero, which is precisely why objective SES shows no visible association in the total sample [12].

**Figure 1:**
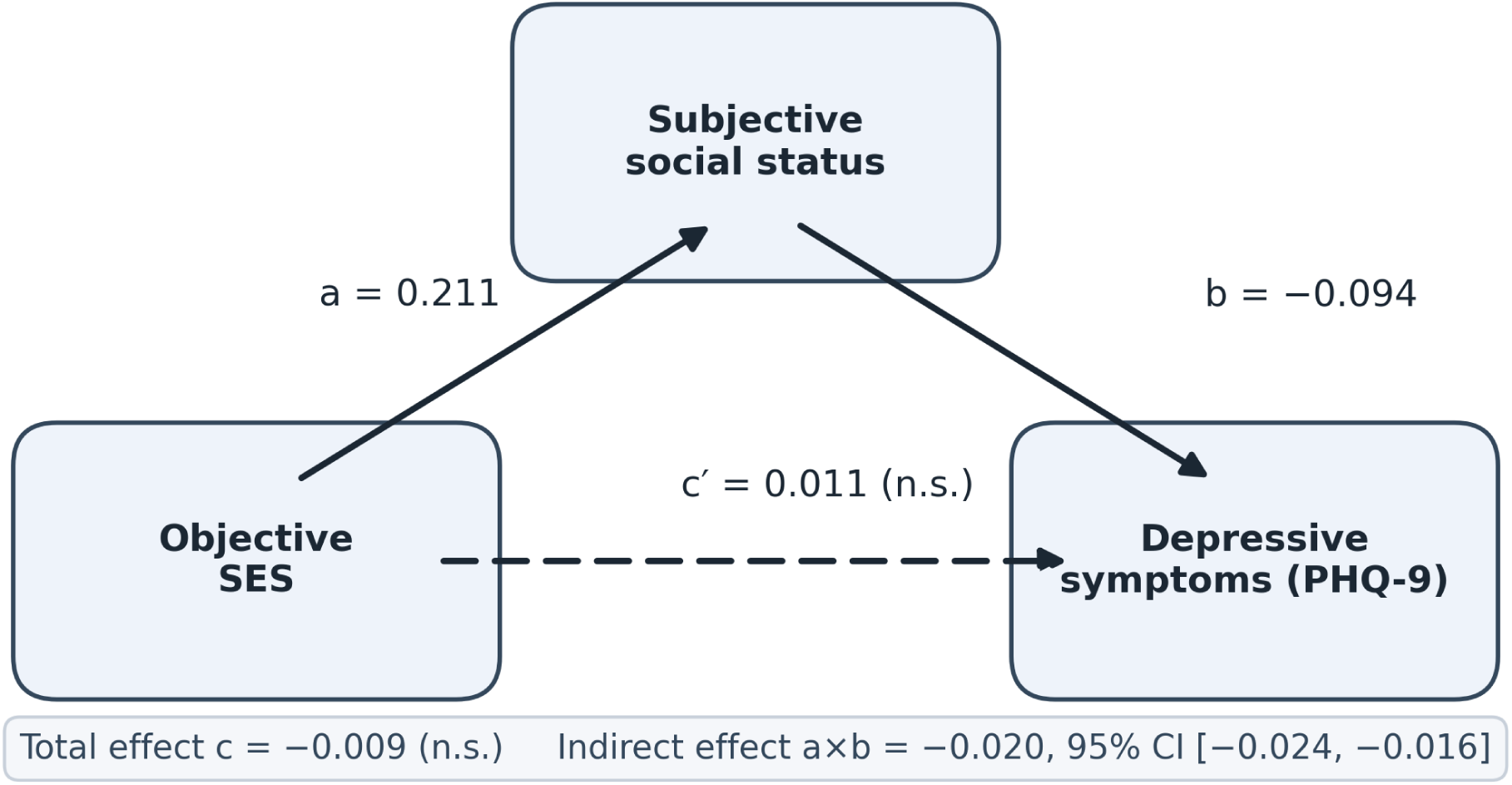
Mediation path diagram. Standardized path estimates from the fully adjusted mediation model (objective SES *−> SSS −> PHQ-9)*.

**Table 2.**
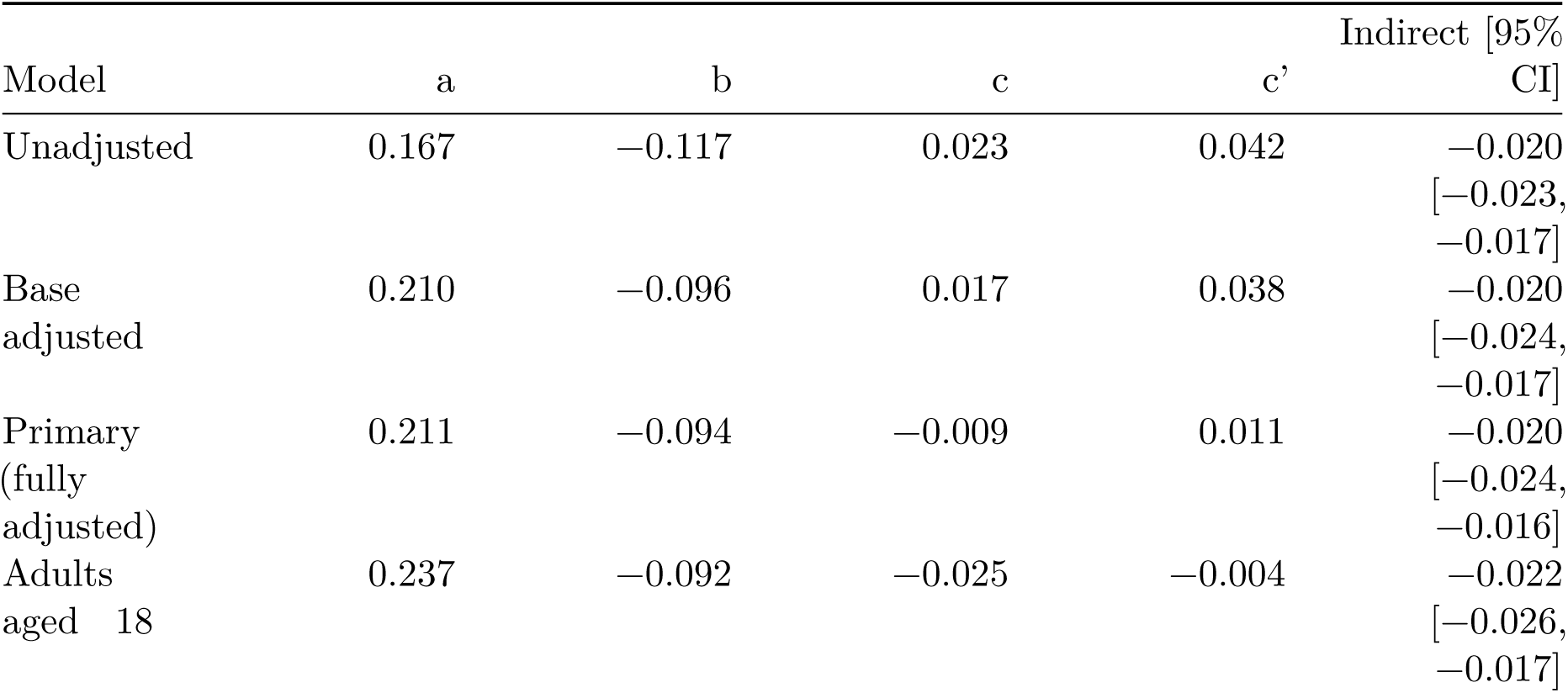
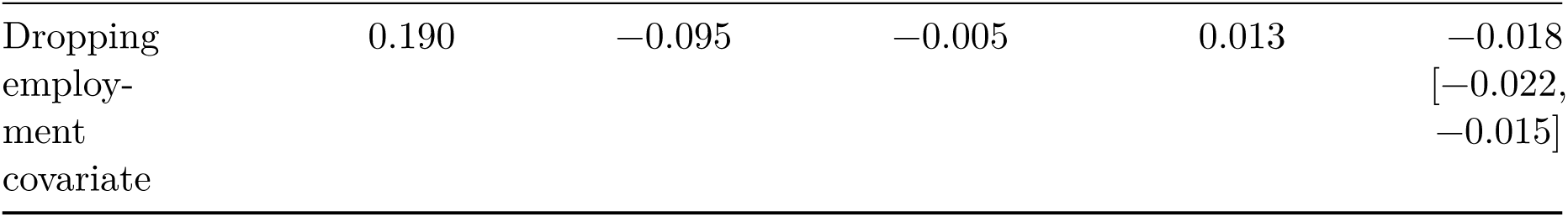
Standardized mediation models (objective SES −> SSS −> PHQ-9). a: objective SES −> SSS; b: SSS −> PHQ-9; c: total effect; c’: direct effect. Indirect effect CI from 3,000 bootstrap resamples. Primary model adjusts for age, sex, marital status, family type, urban/rural residence, employment, chronic disease count, and province fixed effects; “base adjusted” adjusts only for age, sex, marital status, and family type.

Because the total sample includes adolescents, for whom some objective indicators such as employment carry limited meaning, we re-estimated the primary model among adults aged 18 years and older (n = 17,252). Among adults, the total effect turned negative and significant, the direct effect was close to zero, and the indirect effect was again negative and significant, indicating that this association operates almost entirely through subjective social status. The difference between the full sample and adults lies mainly in the direct effect: this effect is nearly zero among adults, so the negative indirect effect shows through in full; the indirect effect itself is of similar magnitude in the two samples, suggesting that the translation of objective conditions into subjective status holds across age groups. Because the direct effect was not significant, this difference should be interpreted with caution; the conclusion remained unchanged after dropping the employment covariate.

### Moderated mediation by marital status, sex, and migration

We further examined whether the indirect effect differs by marital status, sex, and cross-provincial migration status. Table 3 summarizes the differences in indirect effects between strata. Marital status showed the clearest moderation: the indirect effect was stronger among never-married adults than among married adults, and the difference was significant. Because never-married adults tend to be younger, we re-compared the difference within each age band; the differences pointed in the same direction but none reached significance, which may reflect reduced statistical power after stratification or may indicate that the difference itself is not strong. Because only marital status among the three tests reached significance, we treat it as a suggestive finding. Sex and migration status showed no such moderation; both confidence intervals for the differences included zero, so we draw no conclusions about them. The full comparisons within each age band are provided in the Supplementary Material.

**Table 3.** Moderated mediation: bootstrap tests of differences in indirect effects across strata. The significant difference, in bold, is for never-married versus married adults.

| Comparison | Indirect (group 1) | Indirect (group 2) | Difference [95% CI] |
| --- | --- | --- | --- |
| Male versus Female | -0.020 | -0.019 | -0.001 [-0.008, 0.007] |
| Never married versus Married | -0.031 | -0.016 | **-0.015 [-0.026, -0.003]** |
| Local versus Migrant | -0.021 | -0.018 | -0.003 [-0.011, 0.005] |

### Response surface analysis

In the fully adjusted response surface model, the linear term for subjective status (= −0.098) was much larger in absolute value than that for objective status (0.016) and in the opposite direction; the squared term for subjective status was positive (= 0.055), indicating that its negative association with depressive symptoms weakens at higher subjective status. The squared term for objective status and the interaction term were both very small, indicating that the relation between objective status and depression neither shows clear curvature nor works appreciably in concert with subjective status. The model containing only the two status measures explained 2.1% of the total variance in PHQ-9; with covariates added this rose to 9.2% (Figure 2). Overall, subjective social status is more dominant in this set of relations, and the association flattens at higher subjective status, whereas objective status shows no such non-linear feature; full polynomial coefficients are provided in the Supplementary Material.

**Figure 2:**
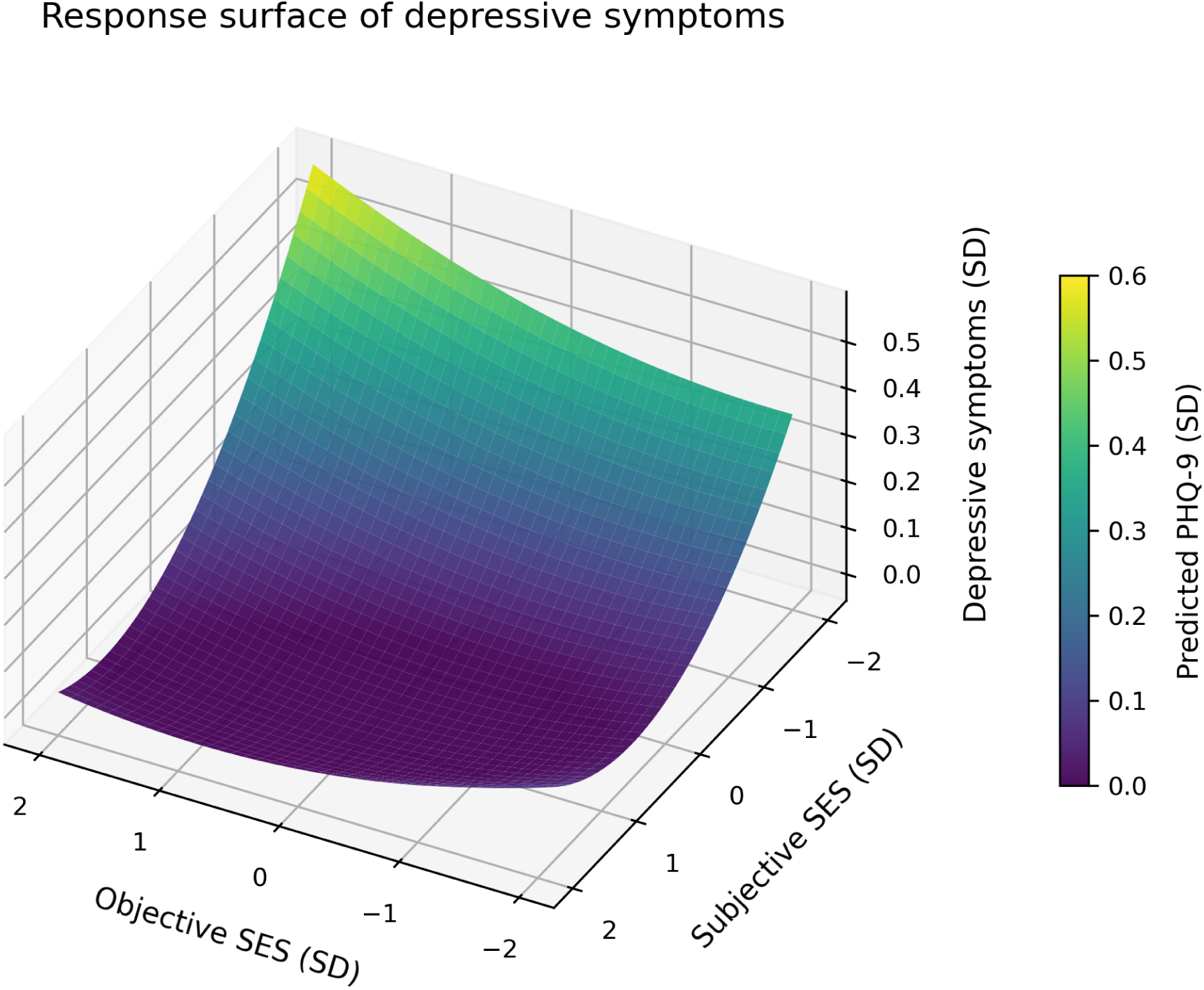
RSA response surface. Response surface for depressive symptoms predicted by objective and subjective SES.

### Sensitivity and robustness analyses

Reversing the direction of mediation produced a non-significant indirect effect, and we found no evidence supporting reverse causation. After excluding participants with PHQ-9 15 or 10, the indirect effect remained significant. Changing the construction of objective SES left the indirect effect almost unchanged, and no confidence interval included zero. To assess unmeasured confounding, we computed E-values: the E-value for the association between subjective status and depressive symptoms was 1.40, a moderately low level, indicating that the robustness of this association to unmeasured confounding is limited [21]; the E-value for the total effect of objective status in the adult sample was even lower. Full results of all sensitivity analyses are provided in the Supplementary Material.

### Mismatch between objective and subjective status and depressive symptoms

To examine whether the state of incongruence between objective conditions and subjective evaluation is itself associated with depressive symptoms, we constructed a mismatch indicator defined as standardized subjective status minus standardized objective status. After grouping by tertiles of the mismatch value, the adjusted depression scores showed a clear gradient: the underestimation group had the highest score, the matched group was intermediate, and the overestimation group had the lowest. The underestimation group had significantly more depressive symptoms than the matched and overestimation groups, while the difference between the overestimation and matched groups was not significant.

In the continuous analysis, because the mismatch value is derived from subjective status minus objective status, entering all three into the same model would result in perfect collinearity; therefore, only the mismatch value and objective status were included. The results showed that, after controlling for objective status, higher mismatch values were associated with fewer depressive symptoms and the association was significant, indicating that when objective conditions are equal, the direction and degree to which an individual’s perception deviates from the objective level is independently associated with depressive symptoms. The product term of objective and subjective status was not significant, indicating that their effects are additive and do not moderate each other. Descriptive statistics of the mismatch indicator and full model results are provided in the Supplementary Material.

## Discussion

### Main findings

In a large national community sample of Chinese residents, subjective social status remained inversely associated with depressive symptoms after accounting for objective SES and a set of covariates. Objective SES showed no clear overall association with depressive symptoms in the total sample, but among adults aged 18 and older it showed a moderate protective association, and this association operated through subjective social status.

In terms of strength, the standardized coefficient for subjective social status was much larger than that for objective status, and the two were in opposite directions. In the adult subsample, about 88% of the total effect of objective status was transmitted through subjective status, and the direct effect was almost negligible; in terms of variance explained, subjective status alone contributed 99% of the incremental variance. In other words, the explanatory role of objective resources for depressive symptoms is almost entirely realized through the pathway of subjective status.

The mismatch analysis further indicated that what matters is not only the level of subjective status but also whether subjective evaluation is consistent with objective conditions: people with adequate objective conditions who rated their status as low had significantly more depressive symptoms, while those who rated themselves higher did not differ significantly from the matched group; after controlling for objective status, the degree of mismatch remained independently associated with depression, and the non-significant product term indicated that the effects of the two are additive.

In the pooled age-adjusted test, the indirect effect was stronger among never-married than married adults, but this difference did not reach significance within age strata, and the indirect effect did not vary by sex or cross-provincial migration. Response surface analysis further showed that depressive symptoms were driven mainly by subjective status, and that the protective effect of subjective status flattened once that status was already high. The difference between the full sample and the adult sample lies mainly in the direct effect: this effect was positive among adolescents and nearly zero among adults, so the indirect effect was no longer offset; the objective status of adolescents derives largely from their parents, and its meaning may differ from that of adults. Because this study was not designed to test this difference, we offer no mechanistic interpretation.

### Comparison with previous studies and mechanistic explanation

Compared with previous studies, this study differs in three respects. First, previous studies mostly reported that depression is more common at lower social status, whereas in the present sample objective SES showed no overall association with depressive symptoms, and its raw correlation with PHQ-9 scores was also nearly zero (r = 0.02). Second, existing research has mostly focused on the separate associations of subjective and objective status with depression and has rarely directly tested whether the state of incongruence between the two is itself associated with depression; our mismatch analysis showed that among people with similar objective conditions, those who rated their status as low had significantly more depressive symptoms. Third, previous tests of group differences mostly relied on informal subgroup comparisons that ignored sampling error; we used moderated mediation analysis to formally test three groups and provided confidence intervals for the differences [12].

How can these differences be explained? We believe they relate to changes in the way Chinese society has been stratified over the past few decades. First, the reference frame for judging one’s own position has changed. With faster population mobility and the spread of social media, the objects of comparison have expanded from neighbors and relatives to a much broader population. A person may be at the top locally in income yet feel left behind after seeing higher-level lifestyles online; subjective evaluation is therefore more likely to deviate from objective circumstances [5]. Second, the faster pace of stratification widens the gap between expected and actual position: some people have seen their objective conditions improve yet still feel insecure; others have stagnant objective conditions but feel stronger frustration because their comparison targets are rising rapidly. This gap itself is associated with negative emotions [22]. Third, subjective status is intertwined with judgments of social fairness. Zhang et al. found in the Chinese working population that about one-third of the effect of subjective status on depression is transmitted through perceived social fairness [6]. The mismatch analysis provides direct empirical support for the above mechanism: if the gap itself did not matter, then people with similar objective conditions should have similar levels of depression regardless of whether they overestimate or underestimate themselves, but the results showed otherwise.

Overall, the direction of this study is consistent with previous research: the association between subjective social status and depressive symptoms is independent of objective SES, a finding reported in both Western and Chinese samples [2,3,6,7], and the association between the subjective-objective status gap and health in East Asia is also supported [8]. The instability of associations between income, education, and depression in Chinese samples [9,10] also explains why the total effect of objective status was not significant in this study.

### Marital status, strengths, and limitations

If the indirect effect is indeed stronger among never-married adults, this finding would agree with an established view: marriage can buffer the link between subjective social status and depressive symptoms. In East Asian settings, marriage is an important marker of adult social standing and provides individuals with an alternative source of identity and social belonging [23]. Never-married adults lack this external identity support and may rely more on occupational achievement and self-evaluation to affirm their self-worth, so subjective social status carries greater psychological weight for them. However, the marital contrast was significant only in the pooled age-adjusted test and not within individual age strata, so it should be regarded as suggestive; this interpretation may also partly reflect the younger age of never-married respondents.

The strengths of this study include its reliance on a large national community sample (N = 19,048) and the inclusion of multiple covariates including province fixed effects; the moderated mediation analysis provided confidence intervals for group differences, avoiding conclusions based solely on subgroup comparisons; and sex-based analyses and reporting followed the Sex and Gender Equity in Research (SAGER) guidelines [20].

There are five main limitations. First, the cross-sectional design cannot support causal inference. The indirect effect is only a statistical decomposition consistent with the hypothesized ordering but insufficient to establish it [24,25]; although the reverse mediation model showed no effect, reverse causality cannot be fully ruled out. Second, subjective and objective status are correlated (r = 0.17), so any statistical decomposition relies on conditional independence assumptions and cannot fully identify their independent effects; the mismatch analysis can only partially alleviate this problem, and it was post hoc and exploratory without correction for multiple comparisons. Third, subjective status was measured with a single item, and common method bias cannot be fully excluded, although Harman’s test did not indicate severe bias [19]; specific values are provided in the Supplementary Material. Fourth, this sample is a non-probability quota sample without sampling weights; the depression detection rate of 21.77% is higher than weighted national estimates [11], should not be taken as population prevalence, and the results are difficult to generalize directly. Fifth, objective status is a composite indicator, and the above conclusion of no association applies only to the operationalization used here; E-value analysis showed that the robustness of the core association to unmeasured confounding is limited, with specific values provided in the Supplementary Material; in addition, the subjective status item was also administered to adolescents aged 12 to 17, and the total effect of objective status reached significance only among adults.

## Conclusion

In a large national community sample of Chinese residents, the association between subjective social status and depressive symptoms was independent of objective SES, and the link between objective status and depression was transmitted almost entirely through subjective status. This indirect association was relatively stronger among never-married adults, but the difference remains suggestive, and no differences were found by sex or cross-provincial migration. These results suggest that depressive symptoms relate not only to how many objective resources a person holds but also to how they evaluate their own social position; therefore, clinical and public health screening should not rely solely on objective indicators such as income and education but should also attend to individuals’ subjective evaluation of their social position. Because of the cross-sectional design, this study cannot support causal inference.

## Data Availability

The data used in this study were derived from the 2022 Psychology and Behavior Investigation of Chinese Residents (PBICR). The datasets are available from the corresponding author on reasonable request with the permission of the PBICR project team.

## Statements and Declarations

### Funding

No funding was received for conducting this study.

### Competing interests

The authors declare that they have no competing interests.

### Ethics approval

The study protocol was approved by the Ethics Committee of the Health Culture Research Center of Shaanxi (approval number JKWH-2022-02). All procedures performed in this study were in accordance with the ethical standards of the institutional research committee and with the 1964 Helsinki declaration and its later amendments.

### Consent to participate

Informed consent was obtained from all individual participants included in the study prior to the survey.

### Data availability

The datasets analysed during the current study are not publicly available due to privacy and ethical restrictions but are available from the corresponding author on reasonable request and with permission of the PBICR project committee. The analysis code is available from the corresponding author on request.

### Authors’ contribution

**Author 1:** Conceptualization, Methodology, Formal analysis, Writing - original draft. **Author 2:** Conceptualization, Supervision, Writing - review & editing.

## Acknowledgements

We thank all participants and the PBICR project committee for their contributions to data collection and survey administration.

---

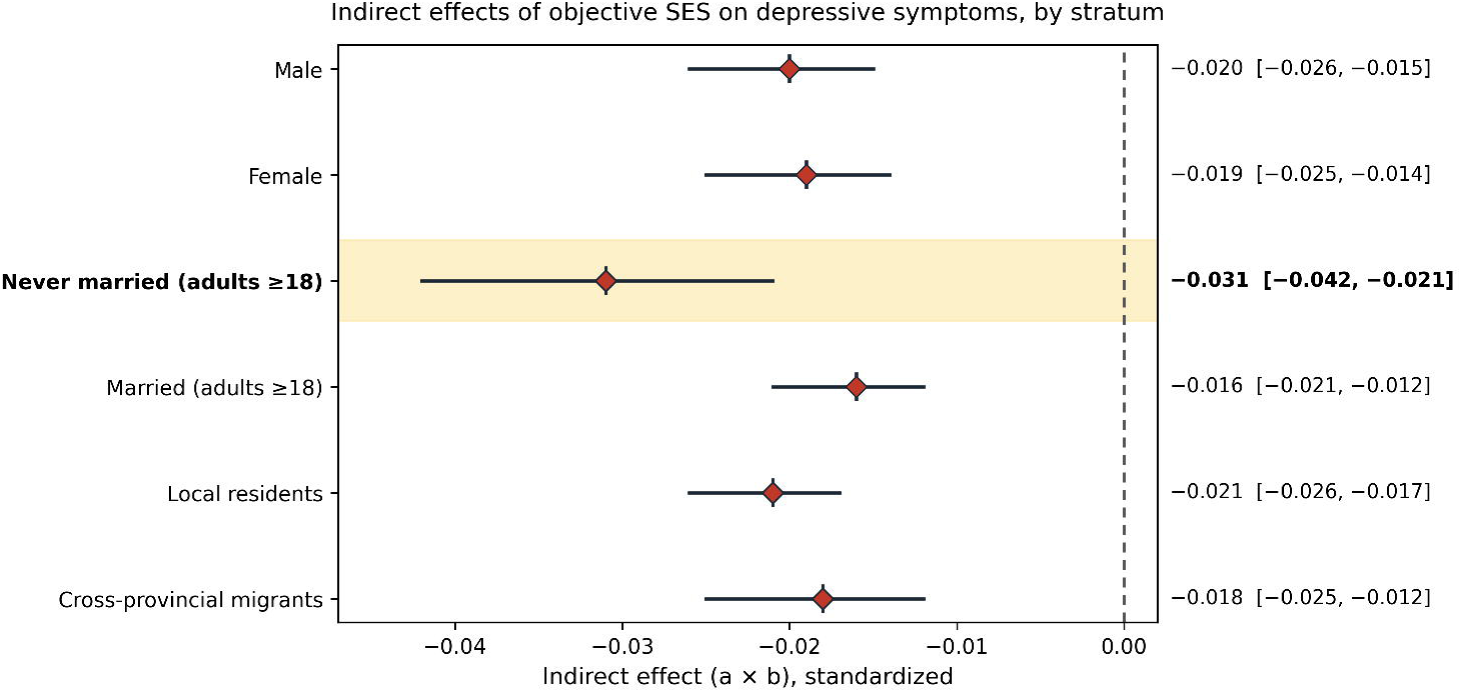

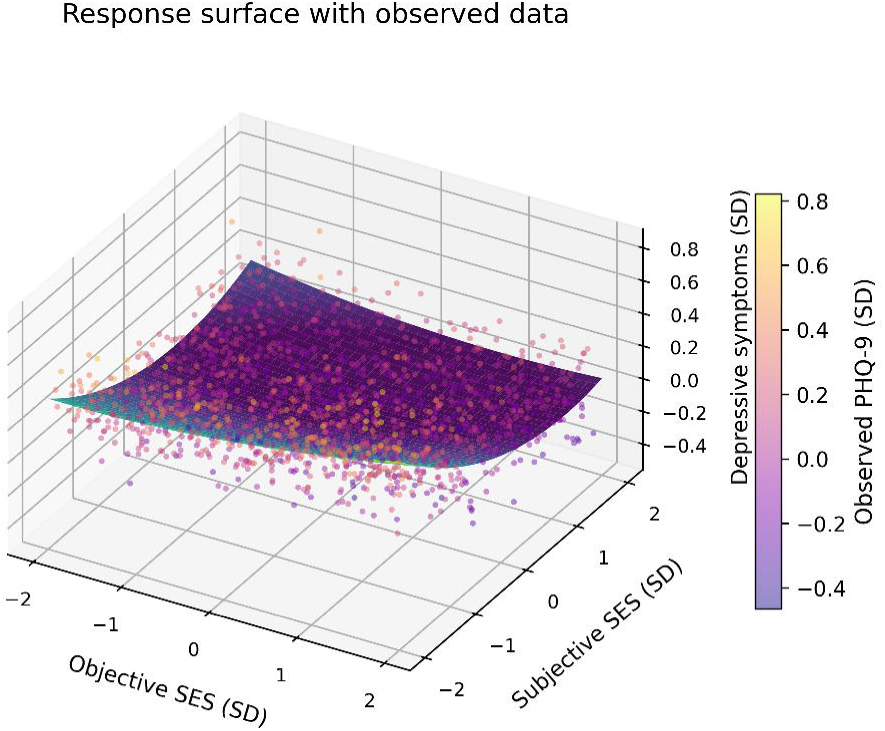

## Notes

### Competing Interest Statement

The authors have declared no competing interest.

### Author Declarations

Ethics committee of the Health Culture Research Center of Shaanxi gave ethical approval for this work (Approval No. JKWH-2022-02). Informed consent was obtained from all participants prior to the survey.

### Summary of Updates

New analysis added: mismatch between objective and subjective status. We constructed an index of status mismatch (standardized subjective social status minus standardized objective socioeconomic status) and examined whether the divergence between objective conditions and subjective appraisal is itself associated with depressive symptoms. Individuals whose objective conditions were not unfavorable but who rated their status low reported markedly more symptoms, and the degree of mismatch remained independently associated with symptoms after objective status was controlled. A note on identification: because the index is defined as the difference between the two status measures, the two cannot enter the same model together, and only objective status was included.

