## supplementary for "Subjective social status is associated with depressive symptoms beyond objective socioeconomic status among Chinese residents: a cross-sectional study"

### Supplementary materials description

These supplementary materials report measurement and validity statistics, stratified and moderated mediation results, sensitivity analyses, and detailed specifications of the statistical analyses that support the primary analyses presented in the manuscript. Unless otherwise noted, all analyses are based on the analytical sample (N = 19,048).

### Figure S1

*Figure S1: Response surface with observed data. The figure displays the polynomial regression surface (objective socioeconomic status, subjective social status, and predicted PHQ-9 scores) obtained from the fully adjusted response surface analysis, with observed data points overlaid. Subjective social status dominated the association with depressive symptoms, and this inverse association attenuated at high levels of subjective status. (Linear coefficient for subjective status = $-0.098$, linear coefficient for objective socioeconomic status = 0.016; squared coefficient for subjective status = 0.055.)*

### Table S1

**Table S1: Measurement and validity statistics.** Correlation coefficients among study constructs and psychometric properties of the PHQ-9.

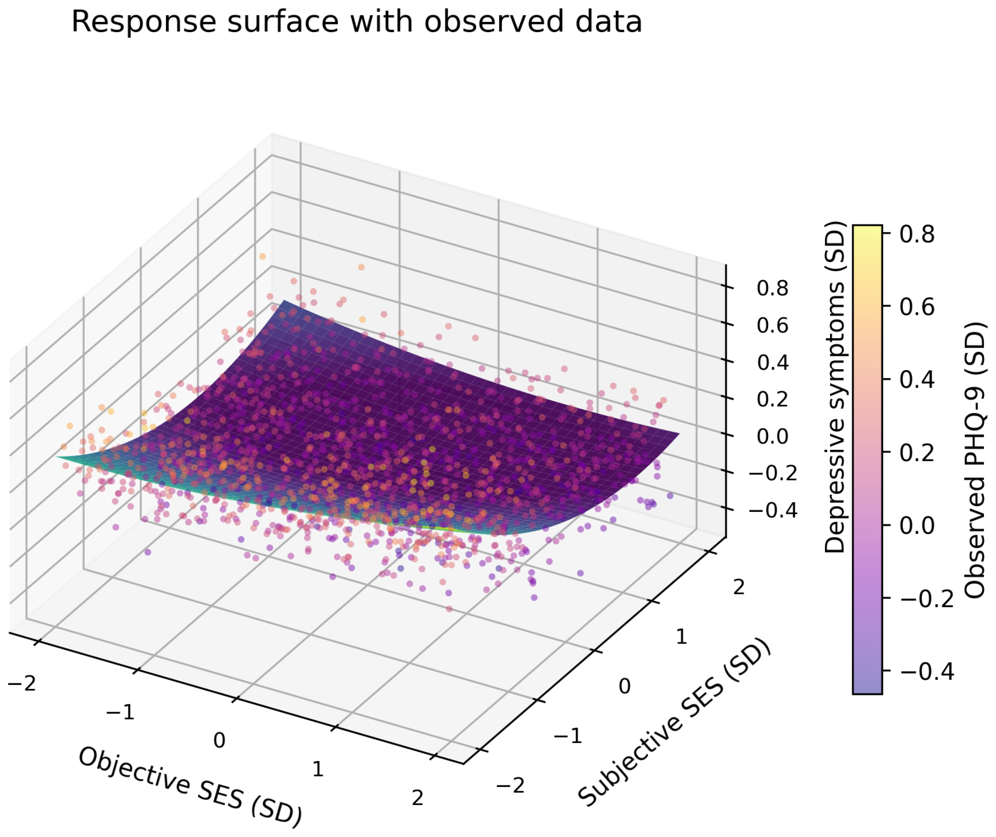


Figure 1: Response surface with observed data

| Statistic | Value | Note |
|---|---|---|
| Correlation between objective socioeconomic status and subjective social status (r) | 0.167 | Weak positive correlation between constructs |
| Correlation between subjective social status and PHQ-9 (r) | −0.110 | Inversely correlated with depressive symptoms |
| Correlation between objective socioeconomic status and PHQ-9 (r) | 0.023 | Negligible association |
| Correlation between income alone and PHQ-9 (r) | 0.002 | No association |
| Cronbach's α (PHQ-9) | 0.912 | Good internal consistency |
| KMO (PHQ-9) | 0.939 | Factorability diagnostic, excellent sampling adequacy |
| Bartlett's test of sphericity (PHQ-9) | $\chi^2$ = 91,054.28, p < .001 | Factorability diagnostic, correlation matrix is not an identity matrix |
| Variance explained by first factor, principal component analysis (PHQ-9) | 59.3% | Consistent with a unidimensional scale |
| Variance explained by first factor, maximum likelihood method (PHQ-9) | 53.8% | Consistent with a unidimensional scale |

*Note: The KMO and Bartlett tests are factorability diagnostics (assessing whether the item correlation matrix is suitable for factor analysis) rather than direct measures of construct validity.*

### Table S1a

**Table S1a: Questionnaire item numbers, original wording, and coding for each measurement indicator.**

| Construct | Item number | Original wording | Response options and coding |
|---|---|---|---|
| Depressive symptoms | Item 119 | Patient Health Questionnaire-9 | 9 items, 2-week recall period, 0 (not at all) to 3 (nearly every day); total score 0-27; scores of 10 or higher defined as probable depression |
| Family per-capita monthly income | Item 84 | Currently, your family's per-capita monthly income is | 10 levels, from below 1,000 yuan to above 15,000 yuan |
| Educational attainment | Item 14 | Your highest level of education (highest degree obtained) | 9 levels, from no formal education to doctoral degree |
| Employment status | Item 11 | Your current employment status | "Employed" coded as employed, remaining options coded as unemployed |
| Housing conditions | Items 81 and 82 | The floor area of your current family residence; the number of properties owned by your family | Standardized and averaged across the two items |
| Subjective social status | Item 76 | How do you perceive the social status of your family | 7-point scale, 1 (very low) to 7 (very high) |

### Table S2

**Table S2: Standardized mediation models stratified by sex, marital status, and migration status (objective socioeconomic status → subjective social status → PHQ-9).** a: objective socioeconomic status → subjective social status; b: subjective social status → PHQ-9; c: total effect; c': direct effect.

| Stratum | n | a | b | c | c' | Indirect effect [95% CI] |
|---|---|---|---|---|---|---|
| Male | 9,313 | 0.206 | −0.098 | 0.019 | 0.039 | −0.020 [−0.026, −0.015] |

| Stratum | n | a | b | c | c’ | Indirect effect [95% CI] |
|---|---|---|---|---|---|---|
| Female | 9,735 | 0.220 | −0.088 | −0.045 | −0.025 | −0.019 [−0.025, −0.014] |
| Never married adults | 5,777 | 0.304 | −0.104 | −0.061 | −0.030 | −0.031 [−0.042, −0.021] |
| Married adults | 10,634 | 0.196 | −0.084 | −0.014 | 0.003 | −0.016 [−0.021, −0.012] |
| Local residents | 13,916 | 0.227 | −0.095 | 0.009 | 0.030 | −0.021 [−0.026, −0.017] |
| Cross-provincial migrants | 5,081 | 0.175 | −0.104 | 0.037 | 0.055 | −0.018 [−0.025, −0.012] |

*Note: Confidence intervals are based on 3,000 percentile bootstrap resamples (random seed 42). Marital stratification is restricted to adults aged 18 years and older; migration stratification uses the full sample.*

### Table S3

**Table S3: Moderated mediation, bootstrap tests of differences in indirect effects.**

| Comparison | Indirect effect (Group 1) | Indirect effect (Group 2) | Difference [95% CI] | Statistically significant |
|---|---|---|---|---|
| Male versus female | −0.020 | −0.019 | −0.001 [−0.008, 0.007] | No |
| Never married versus married adults | −0.031 | −0.016 | −0.015 [−0.026, −0.003] | Yes |
| Local residents versus cross-provincial migrants | −0.021 | −0.018 | −0.003 [−0.011, 0.005] | No |

| Comparison | Indirect effect (Group 1) | Indirect effect (Group 2) | Difference [95% CI] | Statistically significant |
|---|---|---|---|---|
| Never married versus married, ages 18-29 | −0.033 | −0.019 | −0.014 [−0.038, 0.013] | No |
| Never married versus married, ages 30-44 | −0.032 | −0.024 | −0.008 [−0.044, 0.024] | No |
| Never married versus married, ages 45 and older | −0.004 | −0.012 | 0.008 [−0.059, 0.075] | No |

*Note: The difference by marital status was statistically significant in the pooled age-adjusted test, but not statistically significant within any single age stratum (18-29, 30-44, 45 years and older), which is consistent with limited statistical power rather than an absence of effect. Differences by sex and migration status were both not statistically significant.*

### Table S3a

**Table S3a: Full polynomial coefficients of the response surface analysis (fully adjusted model).**

| Term | Coefficient |
|---|---|
| Objective socioeconomic status (linear term) | 0.016 |
| Subjective social status (linear term) | −0.098 |
| Objective socioeconomic status squared term | 0.016 |
| Subjective social status squared term | 0.055 |
| Objective socioeconomic status × subjective social status | −0.021 |
| $R^2$, surface-only model | 2.1% |
| $R^2$, fully adjusted model | 9.2% |

*Note: The linear term for subjective social status is negative and the squared term is positive, indicating that its inverse association with depressive symptoms attenuates at higher levels of subjective status. Both terms for objective status are small, indicating that its relationship with depression displays neither noticeable curvature nor meaningful interaction with subjective status.*

### Table S4

**Table S4: Cutoff sensitivity of the association between objective socioeconomic status and depression (adjusted OR per SD).**

| Cutoff | Prevalence (%) | OR [95% CI] | p |
|---|---|---|---|
| PHQ-9 ≥ 5 | 58.03 | 0.991 [0.956, 1.027] | .62 |
| PHQ-9 ≥ 10 | 21.77 | 0.966 [0.927, 1.007] | .11 |
| PHQ-9 ≥ 15 | 8.35 | 0.989 [0.930, 1.052] | .72 |

*Note: Objective socioeconomic status showed no association with probable depression across any of the cutoff values, consistent with the overall null association observed in the full sample.*

### Table S5

**Table S5: Indirect effects in mediation models under alternative objective socioeconomic status constructions (objective socioeconomic status → subjective social status → PHQ-9).**

| Construction method | Indirect effect [95% CI] |
|---|---|
| Principal component analysis (primary analysis) | −0.020 [−0.024, −0.016] |
| Equal weighting | −0.022 [−0.026, −0.018] |
| Income-dominant | −0.022 [−0.026, −0.018] |

*Note: Indirect effects obtained from the three construction methods were largely identical, and none of the confidence intervals included zero.*

### Table S6

**Table S6: Sensitivity analyses for reverse causality.**

| Model | n | Indirect effect [95% CI] |
|---|---|---|
| Reversed model (objective socioeconomic status → PHQ-9 → subjective social status) | 19,048 | 0.0008 [−0.0009, 0.0025] |
| Excluding PHQ-9 ≥ 15 | 17,457 | −0.016 [−0.019, −0.014] |
| Excluding PHQ-9 ≥ 10 | 14,902 | −0.011 [−0.014, −0.009] |

*Note: The indirect effect in the reversed model was not statistically significant, and the original indirect effect persisted after excluding participants with high scores, providing no evidence to support the reverse pathway.*

### Table S7

**Table S7: Common method bias, Harman's single-factor test.**

| Analysis | Variance explained by the first factor | Note |
|---|---|---|
| PHQ-9 items alone (principal component analysis) | 58.9% | Expected result for a unidimensional scale |
| Subjective social status + PHQ-9 combined (principal component analysis) | 53.1% | No single dominant factor |
| Subjective social status + PHQ-9 combined (maximum likelihood method) | 49.6% | No single dominant factor |

*Note: When analyzing PHQ-9 items alone, the first factor explained 58.9% of the variance, reflecting the unidimensionality of the scale itself; when combined with subjective social status, the variance explained by the first factor did not exceed 53.1%, which does not support the presence of substantial common method bias.*

### Table S8

**Table S8: E-values for unmeasured confounding (VanderWeele & Ding, 2017).**

| Parameter | Estimate | E-value (point estimate) | E-value (confidence interval limit closer to 1) |
|---|---|---|---|
| Path b: subjective social status → PHQ-9 | −0.094 | 1.40 | 1.36 |
| Total effect of objective status in the adult sample | −0.025 | 1.18 | 1.09 |

*Note: The E-value indicates how strong an unmeasured confounder would need to be associated with both the exposure and the outcome to completely explain away the observed association. The E-value for the association between subjective social status and depressive symptoms was 1.40, which falls in the low-to-moderate range, indicating that the robustness of this association to unmeasured confounding is limited. For the small total effect of objective status in the adult sample, the E-value was only 1.18.*

### Table S9

**Table S9: Descriptive statistics for the subjective-objective status mismatch indicator.**

| Statistic | Value |
|---|---|
| Mean | 0.00 |
| Standard deviation | 1.29 |
| Minimum | −4.87 |
| Maximum | 4.55 |
| Correlation with PHQ-9 | −0.10 |
| Number in status underestimation group | 6,356 |
| Number in concordance group | 6,342 |
| Number in status overestimation group | 6,350 |

*Note: The mismatch indicator was calculated by subtracting standardized objective socioeconomic status from standardized subjective social status. Positive values indicate that self-rated status is higher than the position corresponding to objective conditions.*

### Statistical analysis details

- Total PHQ-9 scores were calculated only when at least 7 of the 9 items were completed.
- All continuous variables were standardized to mean 0 and standard deviation 1 prior to analysis.
- Bootstrap confidence intervals for indirect effects were based on 3,000 percentile resamples with the random seed fixed at 42.
- The fully adjusted (primary) model controlled for age, sex, marital status, family type, urban-rural residence, employment status, number of chronic conditions, and province fixed effects (top 20 provinces plus “Other”).
- Migration-stratified models and moderated mediation models did not include province fixed effects to ensure adequate sample sizes within each stratum.
- Employment status served both as a constituent component of the objective socioeconomic status index and as a covariate in the models, introducing the possibility of over-adjustment;

therefore, an additional sensitivity analysis was conducted removing the employment co-variate. After removal, the null association for objective socioeconomic status remained unchanged (adjusted total effect = −0.005, $p = .56$), and the indirect effect remained statistically significant (−0.018 [−0.022, −0.015]).
